# Transcranial sonography reveals striatal neurodegeneration in female XDP-causing variant carriers

**DOI:** 10.64898/2026.05.27.26354192

**Authors:** Martje G. Pauly, Cid Czarina E Diesta, Paulo Cataniag, Max Borsche, Jed Ong, Teresa Kleinz, Jan Uter, Jean Quint L. Oropilla, Max Brand, Shela Marie Algodon, Christine Klein, Ana Westenberger, Norbert Brüggemann

## Abstract

**Objectives:** X-linked dystonia-parkinsonism is a neurodegenerative movement disorder with predominant striatal pathology in affected males, who frequently show hyperechogenicity of the lentiform nucleus on transcranial sonography. We aim to investigate female mutation carriers and female healthy controls using transcranial sonography to identify potential abnormalities in the striatum, substantia nigra, and ventricular system.

**Methods:** We examined 81 participants (35 female mutation carriers and 46 female controls) using transcranial sonography to assess the presence of hyperechogenicity of the lentiform nucleus, the area of substantia nigra hyperechogenicity, and the widths of the lateral and third ventricles. Clinical evaluation focused on dystonic and parkinsonian symptoms, and we determined genotypes relevant for four X-linked dystonia-parkinsonism genetic modifiers.

**Results:** Female mutation carriers showed more subtle parkinsonian signs compared with controls. The prevalence of hyperechogenicity of the lentiform nucleus was higher in female mutation carriers and was associated with a more unfavorable genetic modifier profile. No relevant abnormalities were observed in the substantia nigra or the ventricular system. Imbalanced X-chromosome inactivation in favor of the wildtype allele expression was not significantly associated with clinical severity or hyperechogenicity of the lentiform nucleus frequency, although female mutation carriers with such an imbalance showed no parkinsonian signs and only rarely hyperechogenicity of the lentiform nucleus (1/8, 13%).

**Conclusions:** Women carrying the X-linked dystonia-parkinsonism-causing variant display subtle parkinsonian signs and frequently exhibit hyperechogenicity of the lentiform nucleus, supporting hyperechogenicity of the lentiform nucleus as a sensitive imaging marker of early neurodegenerative change, especially in those with higher genetic risk.

**Summary for Social Media If Published:**

1. What is the current knowledge on the topic? Hyperechogenicity of the lentiform nucleus as part of the striatum has not only been found in patients with X-linked dystonia-parkinsonism but also in variant carriers prior to first symptoms.
2. What questions did this study address? Do women with the X-linked dystonia-parkinsonism causing variant exhibit hyperechogenicity of the lentiform nucleus, and do they show subtle signs without full manifestation of the disorder?
3. What does this study add to our knowledge? Women carrying the X-linked dystonia-parkinsonism-causing variant show pathology of the striatum even without manifesting symptoms, but have more subtle parkinsonian signs than healthy controls.
4. How might this potentially impact the practice of neurology? Women carrying the X-linked dystonia-parkinsonism-causing variant should be evaluated and monitored for pathology in the striatum and development of parkinsonian signs.

## Introduction

Women carrying the X⍰linked dystonia⍰parkinsonism (XDP)-causing pathogenic variant^1–3^ are usually considered clinically unaffected, yet accumulating evidence suggests that neurodegenerative changes may already be present in non-manifesting male carriers long before overt motor symptoms emerge^4,5^. In male XDP patients and non⍰manifesting carriers, hyperechogenicity of the lentiform nucleus (LN) on transcranial sonography (TCS) has been established as a sensitive marker of striatal pathology and is associated with a genetic modifier profile linked to earlier age at onset and more pronounced neurodegeneration^6^. Whether female mutation carriers share this subclinical striatal phenotype, and to what extent it translates into subtle parkinsonian signs, has remained unknown.

This question is particularly relevant for women of Filipino descent, in whom XDP is endemic and where parkinsonism appears to be more prevalent among women than men^7^, raising the possibility that XDP⍰related parkinsonism in female carriers is underrecognized. At the same time, recent functional imaging work in asymptomatic female mutation carriers has pointed to altered neuroenergetics, hinting at early brain involvement despite a lack of manifest dystonia or parkinsonism^8^.

Previous studies identified four genetic modifiers—a (CCCTCT) repeat expansion within the SVA (short interspersed nuclear element [SINE]–variable number tandem repeat [VNTR]–Alu) retrotransposon insertion and three modifiers tagged by single-nucleotide polymorphisms (SNPs) —that significantly influence age at onset (AAO) in XDP^1,9^. Variation in the DNA repair gene *MSH3* also affects AAO and repeat stability^9,10^. These modifiers enable estimation of expected AAO in male non-manifesting carriers, in which caudate nucleus volume correlates with time to onset, suggesting a direct role in neurodegeneration^4^.

X-chromosome inactivation (XCI) is usually random and expression of the wild-type and mutant X chromosome is balanced in most women. In some cases, however, inactivation of the wild-type X chromosome exceeds 70%, leading to marked skewing and predominant expression of the mutant allele. This imbalanced or “skewed” XCI is considered a main mechanism that may contribute to phenotypic expression in heterozygous women is skewed XCI^11,12^.

Against this background, we used TCS and detailed clinical assessments to test whether asymptomatic female XDP mutation carriers (fMC) already show LN hyperechogenicity and subtle motor changes, and how these relate to their genetic modifier profile and X⍰chromosome inactivation status, by investigating 81 women of Filipino descent, 35 of them with a genetically confirmed XDP-causing SVA retrotransposon insertion.

## Study participants and methods

### Participants

Female participants of Filipino descent, mostly relatives of patients with XDP, were included as part of multimodal studies either at the University of Lübeck, Germany, from 2013 to 2022 or at Makati Medical Center, Manila, Philippines, from 2023 to 2024. All 81 participants showed no abnormalities upon neurological examination and had no diagnoses of a movement disorder. Of the 81 participants, 35 were genetically confirmed heterozygous carries of the SVA retrotransposon insertion in *TAF1* and were therefore classified as fMC. Before participation, all participants were informed about the study in Tagalog, the official language of the Philippines, and informed written consent was obtained. The study was approved by both local ethics committees of the University of Lübeck and the Makati Medical Center. The study was conducted in accordance with the Declaration of Helsinki.

### Clinical examination

Clinical examinations were performed by movement disorder specialists, who were blinded with regard to the genetic status. The Burke-Fahn-Marsden Dystonia Rating Scale^13^ (BFMDRS) was used to assess signs of dystonia. In addition, in participants investigated in 2024, the part III of the Movement Disorder Society-sponsored revision of the Unified Parkinson’s Disease Rating Scale (MDS-UPDRS)^14^ was used to assess parkinsonian motor signs, and the XDP score of the Movement Disorder Society of the Philippines (XDP-MDSP-RS)^15^ was used to assess dystonic symptoms (part I), parkinsonian symptoms (part II), non-motor features (part III), and activities of daily life (part IV). Part V (global impression) was not used for analysis in this study.

### Genetic testing

All participants underwent genetic testing for the presence of the SVA retrotransposon insertion in *TAF1*^*2,16*^. Furthermore, the number of (CCCTCT) repeats (RN)^1,3^ was determined by PCR-based fragment analysis. The genotypes at the three SNPs tagging the genetic modifiers of AAO in XDP (rs245013, rs33003, rs4724769)^9^ were determined by Sanger sequencing. In line with previous work^9^, the calculated AAO (cAAO) was derived from the RN and genotypes of the three SNPs. The calculated disease duration (cDD) was defined as the time in years between cAAO and age at examination (AAE)^11,17^.

### Transcranial sonography

TCS was carried out by the same examiner (N.B.) in all participants. Between 2013 and 2014, TCS was performed using a Siemens Acuson Antares system, and since then, with an Esaote Mylab alpha, each equipped with a 1-4 MHz sector probe, following a previously established protocol^6,18^.

At the level of the basal ganglia, we refer to the LN because the pallidum and putamen cannot be reliably differentiated with the spatial resolution in TCS. On both sides, LN echogenicity was assessed qualitatively. If either side demonstrated a higher echogenicity than the surrounding white matter, hyperechogenicity of the LN (LN+) was rated as present, if no hyperechogenicity was observed on both sides, it was rated as absent (LN-).

The midbrain was investigated in the axial plane via the temporal bone window. The midbrain was identified by its characteristic butterfly shape, with the cerebral peduncles forming the “wings” and the tegmentum in the center. SN appears as a hyperechogenic (bright) region (SN+) within the ventral midbrain, and planimetric measurements were applied to quantify its size. Both right and left SN+ were measured in cm^2^, and the maximum value was used for further analysis as SN+_max_ . SN+ _max_ was considered pathological if it exceeded 0.20 cm^2^.

The width of the third and lateral ventricles was measured at the thalamus level in mm. For the lateral ventricles, both right and left cella media (pars centralis) of the lateral ventricles were assessed and the mean of both measurements was calculated.

### X-chromosome inactivation

The degree of XCI balance or imbalance was assessed using a standard method, i.e., by HpaII restriction enzyme digestion of the AR gene, exploiting a polymorphic CAG repeat near the HpaII site in exon 1 to distinguish between alleles in women as previously described^17,19^. In samples showing XCI imbalance (i.e., XCI ratios between 0:100 and 35:65), we sequenced the cDNA region spanning the only variant constituting the XDP-specific haplotype that was found to be transcribed - disease-specific variant 3 [DSC3] at chrX:71529785C>T per GRCh38. This allowed us to determine the imbalance direction: If the sequencing electropherogram indicated a higher T peak height at the DSC3 position, it was interpreted as XCI skewing toward preferential expression of the allele harboring the SVA insertion; conversely, a higher C peak was interpreted as the wild-type allele being preferentially expressed. Blood RNA was collected in PAXgene tubes (PreAnalytiX, Qiagen/BD), extracted according to the manufacturer’s protocol, and reverse-transcribed into complementary DNA (cDNA) using a standard kit (Fermentas, Thermo Fisher Scientific). Sanger sequencing of cDNA segments was performed on an ABI 3500XL Genetic Analyzer (Applied Biosystems).

### Statistical Analysis

Statistical analysis of data was performed using SPSS (Version 29.0.2.0) and Graphs were created using Microsoft Excel (Version 16.93.1). The Shapiro-Wilk test was used to test for normality of data. Comparison of groups was performed by independent t-test if normality was confirmed or non-parametric t-test if not. Chi-square test was used to compare frequencies. Analysis of Covariance (ANCOVA) or Quade nonparametric ANCOVA were performed for the width of the ventricular system and MDS-UPDRS part III to correct for different AAE in the groups. Pearson correlation or partial correlation to correct for AAE was performed to test for correlation between variables. A p-value < 0.05 was considered to be significant.

## Results

### Demographic and clinical data

We included 35 fMC and 46 female HC in our study. The AAE was higher in the fMC group than in the HC group (p<0.001, Table 1) which was taken into consideration for further analyses. Clinical data was available for 33 fMC (94%) and 35 HC (76%) for BFMDRS, for 17 fMC (49%) and 25 HC (54%) for the MDS-UPDRS part III, and for 16 fMC (46%) and 24 HC (52%) for XDP-MDSP-RS. There was no difference in the clinical scores between fMC and HC for BFMDRS and XDP-MDSP-RS. The score of the MDS-UPDRS part III was higher in fMC (4.8±5.7) compared to HCs (1.9±5.0, p-value = 0.016) (Fig. 2A-C, Table 1). However, MDS-UPDRS III correlated with AAE in fMC (rho = 0.670, p-value = 0.003) and in HC (rho = 0.527, p-value=0.007), and the difference between groups was not significant if corrected for AAE (p-value = 0.305). MDS-UPDRS part III did not correlate with cDD corrected for AAE (rho = 0.300, p-value = 0.399).

**Table 1.** Demographic, clinical, and sonography data.

|  | **fMC**<br>**N = 35** | **HC**<br>**n = 46** | **p-values\*** |
| --- | --- | --- | --- |
| **Demographic information** |  |  |  |
| AAE (y) | 52.0 (13.25) | 41.7 (11.1) | < 0.001 |
| eAAO (y) | 39.75 (6.03) | - | - |
| eDD (y) | 12.23 (14.40) | - | - |
| **Clinical information** |  |  |  |
| XDP-MDSP-RS | 5.8 (5.4) | 5.1 (5.5) | 0.692 |
| MDS-UPDRS III | 4.8 (5.7) | 1.9 (5.0) | 0.016 |
| BFMDRS | 1.0 (1.6) | 1.0 (1.4) | 0.528 |
| **Sonography** |  |  |  |
| LN+ | 12/30 | 6/43 | 0.011 |
| SN+max (cm<sup>2</sup>) | 0.08 (0.02) | 0.08 (0.03) | 0.552 |
| Third ventricle (mm) | 3.4 (1.6) | 2.2 (1.0) | 0.061 |
| Lateral ventricle (mm) | 14.9 (3.3) | 15.3 (2.0) | 0.512 |

Parenthesis: standard deviation; p-values given for HC and fMC. fMC: female mutation carrier:
HC: healthy controls; AAE: age at examination; y: years; cAAO: calculated age at onset; cDD:
calculated disease duration; BFMDRS motor: motor part of the Burke-Fahn-Marsden Dystonia
Rating Scale; MDS-UPDRS III: Movement Disorder Society-sponsored revision of the Unified
Parkinson's Disease Rating Scale Part III; XDP-MDSP-RS: XDP Movement Disorder Society of the
Philippines score. LN+: Hyperechogenicity of the lentiform nucleus; number after / indicates n
for the specific variable, SN+<sub>max</sub>: maximum size of hyperechogenicity of the substantia nigra.

**Figure 1:**
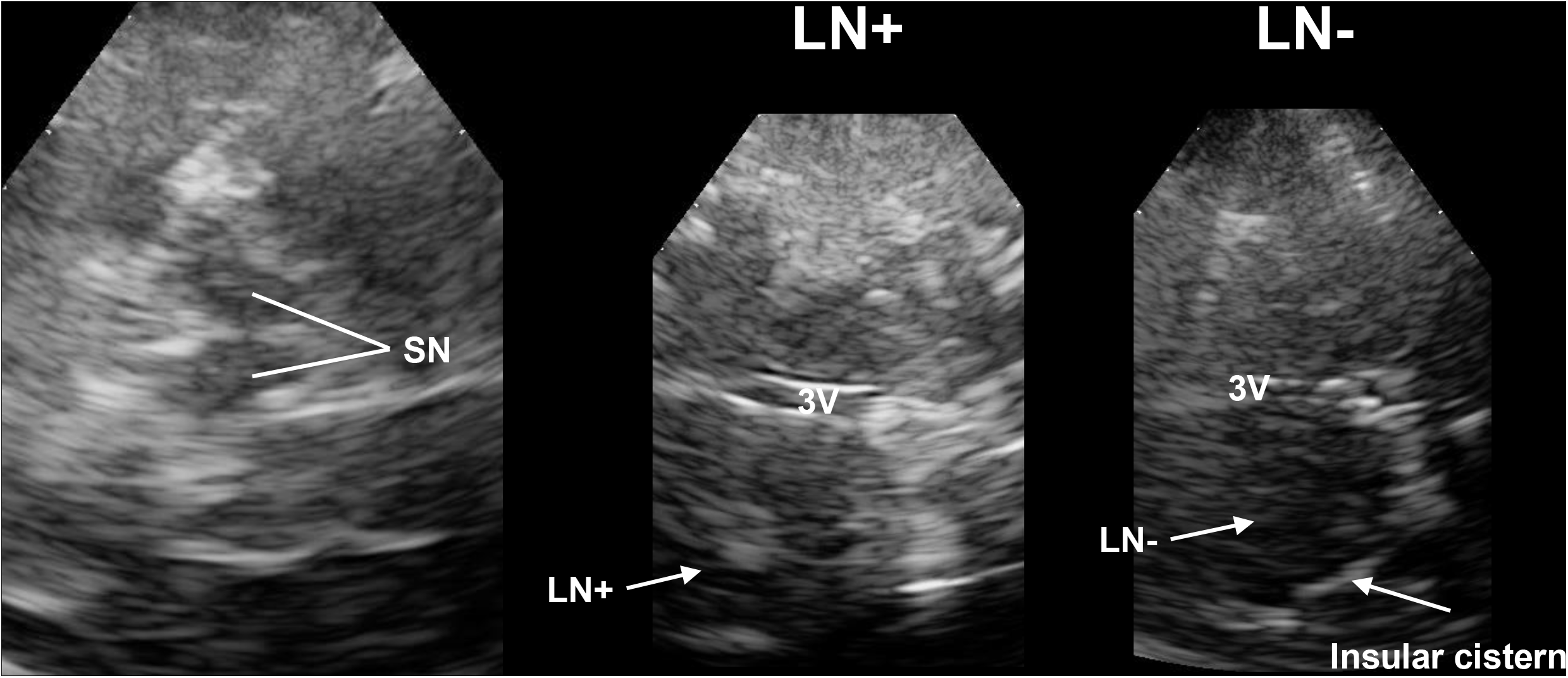
Transcranial sonography. Transcranial sonography imaging of the substantia nigra (SN), the third ventricle (3V), the lentiform nucleus with (LN+) or without hyperechogenicity (LN-) in female mutation carriers (fMC).

**Figure 2:**
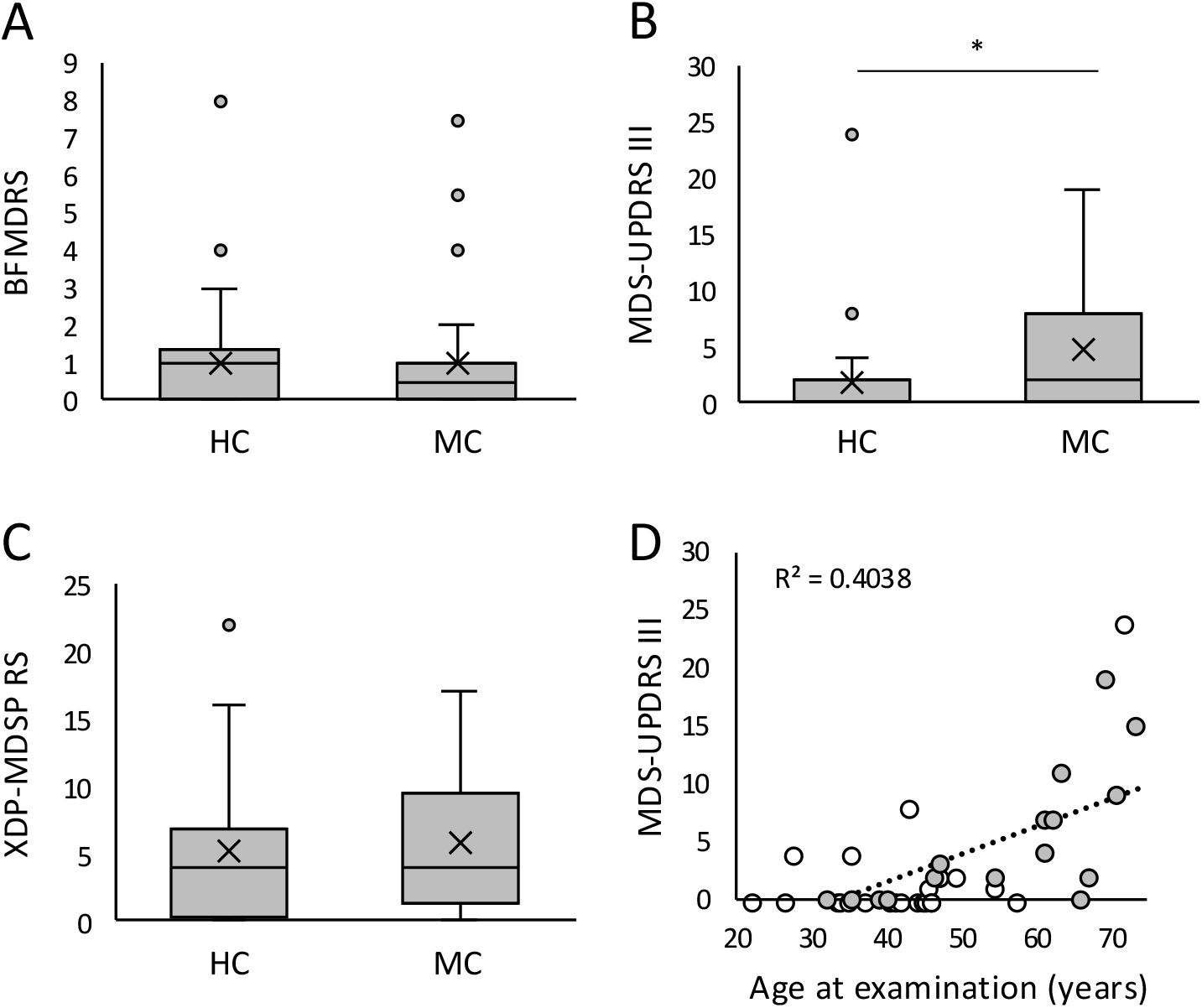
Clinical scores. Clinical scores in regard to the group showing the 25th to 75th percentile as a box, the median as a line, the mean as an X, the minimum and maximum value in the range of 1.5x interquartile range as whiskers, and values outside of the 1.5x interquartile range as dots. A) The motor part of the Burke-Fahn-Marsden Dystonia Rating Scale (BFMDRS) of healthy controls (HC) and mutation carriers (MC); B) The Movement Disorder Society-sponsored revision of the Unified Parkinson’s Disease Rating Scale Part III (MDS-UPDRS III) for HC and MC; C) The X-linked Dystonia-Parkinsonism (XDP) Movement Disorder Society of the Philippines Rating Scale (XDP-MDSP-RS) of HC and MC. D) Correlation between age at examination and MDS-UPDRS III for HC (white dots) and MC (gray dots). *: p-value < 0.05; Line: trend line.

## Transcranial sonography

### Temporal bone window and data availability

All but one fMC had an adequate temporal bone window to assess at least one of the TCS parameters. Data on LN+ were available for 30 fMC (86%) and 43 HC (94%). Data on SN+_max_ were available for 28 fMC (80%) and 44 HC (96%), on third ventricle width in 34 fMC (97%) and 46 HC (100%), and on lateral ventricle width in 27 fMC (77%) and 35 HC (67%).

### Hyperechogenicity of the lentiform nucleus

In the fMC group, 12 out of 30 participants (40%) showed LN+, compared with 6 out of 43 participants (14%) in the HC group (Pearson Chi-Square = 6.453, p-value = 0.011) (Fig.3A). There were no differences in clinical scores in the overall comparison of participants with LN+ and LN-regardless of the genetic status (BFMDRS p-value = 0.713, MDS-UPDRS III p-value = 0.234, XDP-MDSP-RS p-value = 0.667) nor in the fMC group (BFMDRS p-value = 0.819, MDS-UPDRS III p-value = 0.438, XDP-MDSP-RS p-value = 0.833) and in the HC group (BFMDRS p-value = 0.760, MDS-UPDRS III p-value = 0.737, XDP-MDSP-RS p-value = 0.366). The cDD was significantly longer in fMC with LN+ (19.9±11.8 years) than in those without LN+ (5.9±13.9 years, p-value = 0.010) (Fig. 3B). While AAE was higher in fMC with LN+ compared to LN-(58.7±8.1 years vs. 44.6±12.0 years, p-value = 0.001), there was no difference between AAE between LN+ and LN-in the HC group (45.5±6.2 years vs. 40.1±10.4 years, p-value = 0.223). There was also no difference in RN between LN+ and LN-(43.9±3.7 vs. 43.5±3.2, p-value = 0.770) (Fig. 3C).

**Figure 3:**
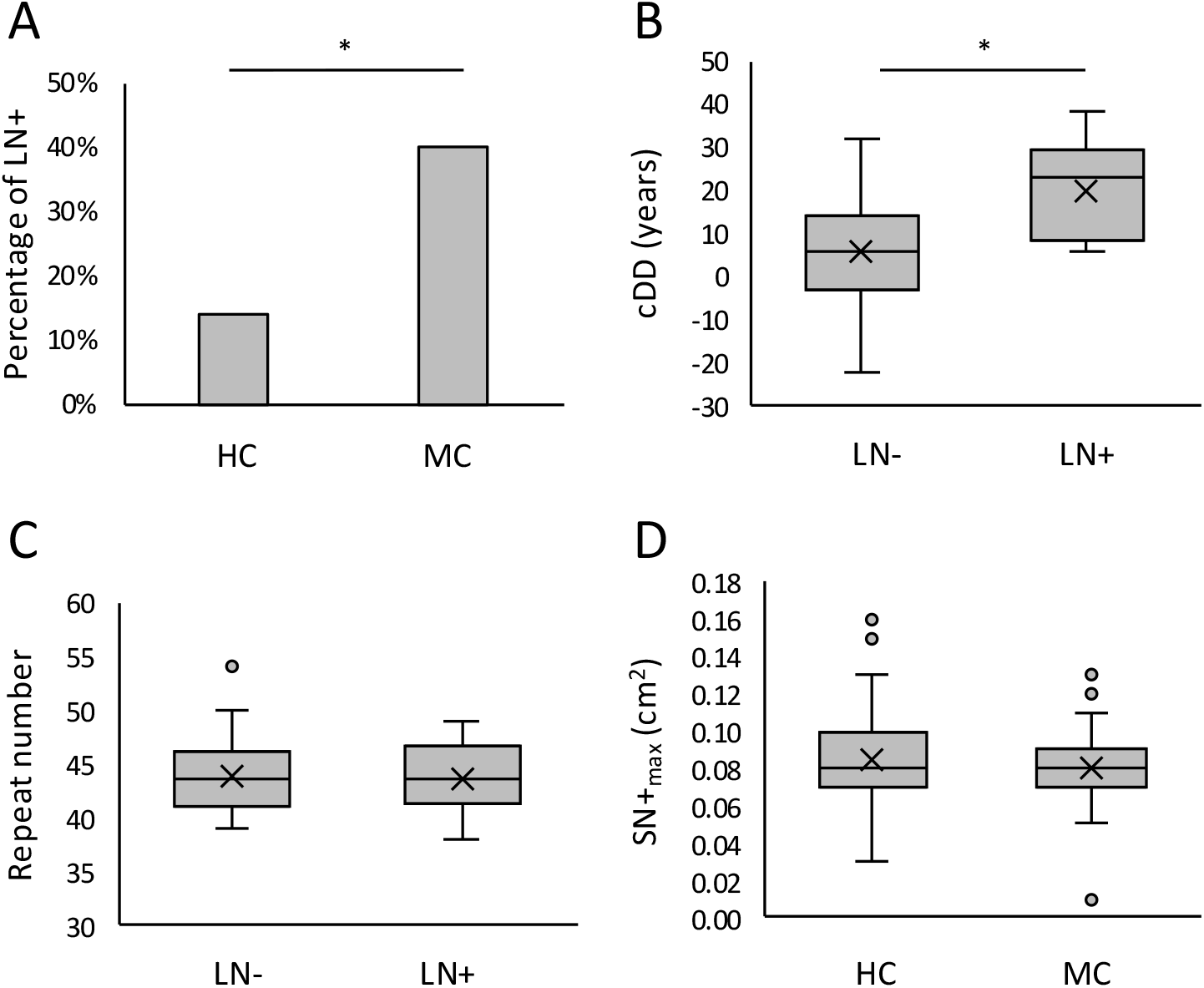
Hyperechogenicity of the lentiform nucleus. A) Percentage of participants with hyperechogenicity of the lentiform nucleus (LN+) in the group of healthy controls (HC) and mutation carrier (MC); B) calculated disease duration (cDD) in the group with LN+ and without hyperechogenicity (LN-); C) repeat number in LN+ and LN-; D) maximum size of in substantia nigra (SN+_max_) in cm^2^ in regard to the group showing the 25th to 75th percentile as a box, the median as a line, the mean as an X, the minimum and maximum value in the range of 1.5x interquartile range as whiskers, and values outside of the 1.5x interquartile range as dots. *: p-value < 0.05.

### Hyperechogenicity of the substantia nigra

SN+_max_ did not differ between groups (p-value = 0.552) and was 0.08±0.02cm^2^ (0.01 – 0.13cm^2^) in fMC and 0.08±0.03cm^2^ (range 0.03 – 0.16cm^2^) in HC (Fig. 3D). None of the participants had an SN+_max_ exceeding 0.20 cm^2^.

### Ventricular system

The third ventricle tended to be wider in fMC (3.4±1.6mm) than in HC (2.2±1.0mm), although this difference did not reach statistical significance (p-value=0.061). There was no significant group difference in lateral ventricle width (fMC: 14.9±3.3mm; HC: 15.3±2.0mm, p=0.512). The AAE correlated with both lateral ventricle width (rho = 0.258, p-value = 0.043) and third ventricle width (rho = 0.671, p-value <0.001) (Fig.4C+D).

**Figure 4:**
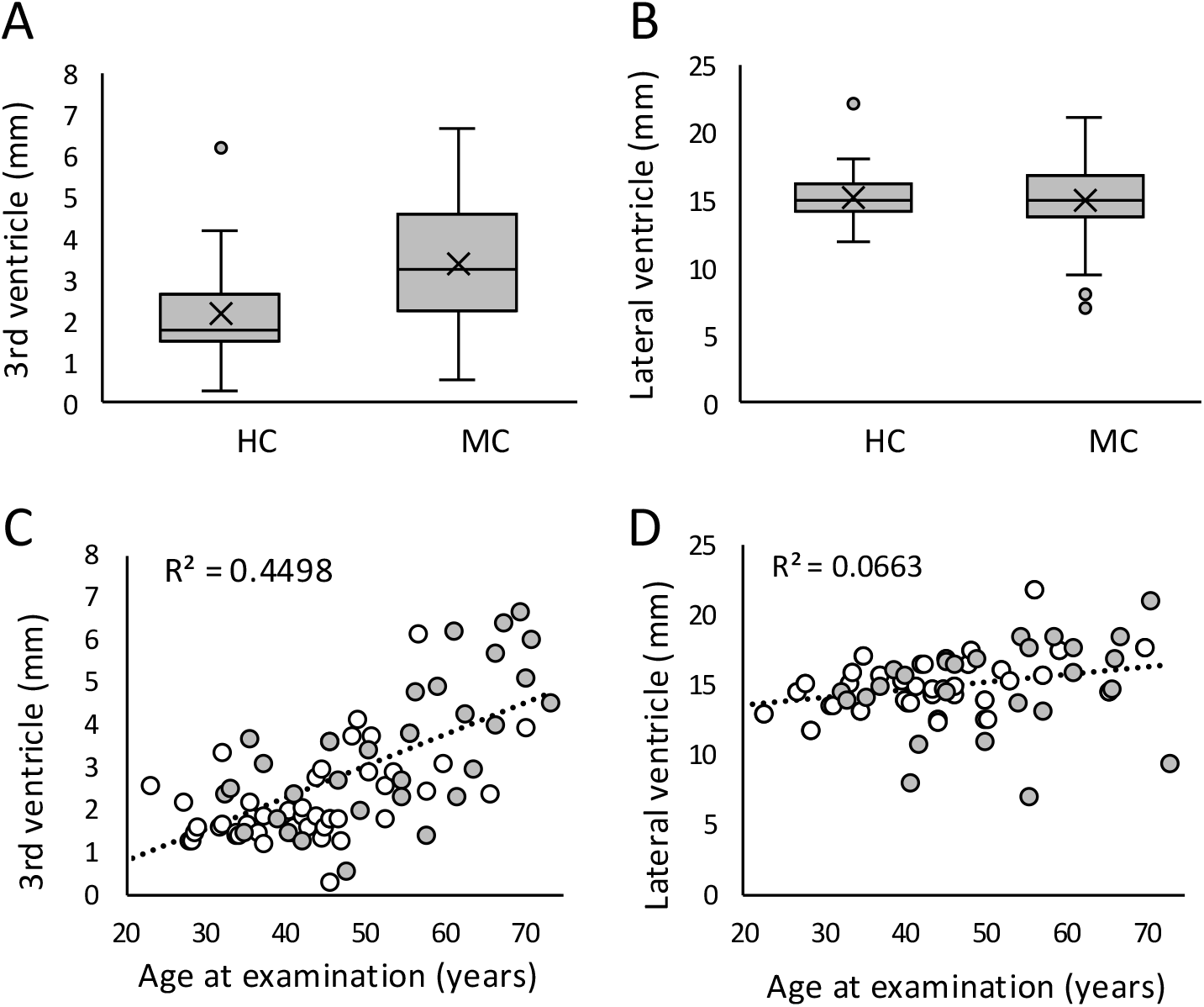
Third and lateral third ventricles. Width of third ventricle and lateral ventricle in regards to the group showing the 25th to 75th percentile as a box, the median as a line, the mean as an X, the minimum and maximum value in the range of 1.5x interquartile range as whiskers, and values outside of the 1.5x interquartile range as dots. A) The width of the third ventricle in mm of healthy controls (HC) and mutation carriers (MC) B) the width of the lateral ventricles of HC and MC; Correlation between C) the width of the third ventricle and age at examination and D) the width of the lateral ventricles and age at examination for HC (white dots) and MC (gray dots). Line: trend line.

### X-chromosome inactivation

Information on XCI was available for all 35 fMC (100%). XCI was balanced in 23 fMC (66%), skewed in favor of the wildtype allele in nine fMC (26%), and skewed in favor of the mutated allele in three fMC (8%). There was no difference between these three groups (balanced, skewed_wt, skewed_mut) in regard of the BFMDRS (balanced: 1.04±0.39, skewed_wt: 0.94±0.42, skewed_mut: 0.33±0.33, p-value=0.690), the MDS-UPDRS III (balanced: 4.57±1.64, skewed_wt: 0.00±0.00, skewed_mut: 2.00±1.56, p-value=0.538), and the XDP-MDSP-RS (balanced: 6.71±2.14, skewed_wt: 4.33±1.76, skewed_mut: 1.67±0.88, p-value=0.367). Of the fMC with information available for LN+, there were 10 out of 19 fMC (53%) in the balanced group, one out of eight (13%) in the skewed_wt group, and one out of three (33%) in the skewed_mut group. The frequency of LN+ did not differ between groups (p-value=0.147).

## Discussion

In this study, we show that female carriers of the XDP-causing mutation (i) show no more subtle parkinsonian signs compared with female HC, (ii) have a higher prevalence of LN+, which is associated with a longer estimated disease duration, and (iii) do not display ultrasound abnormalities of the SN or ventricular system.

While the pathophysiology mainly affects the striatum, as shown in neuropathological and imaging studies^20–29^, more recent work in a mouse model also demonstrates white matter involvement characterized by oligodendrocyte dysfunction and myelin loss^30^. In regard to treatment, deep brain stimulation of the pallidum provides substantial symptomatic benefit^31^, but no causative or disease-modifying treatment is currently available. However, recent preclinical studies have reported promising advances in the development of gene-modifying treatment strategies in mouse^32^ and cell^33^ models.

Although XDP usually affects men because of its X-linked inheritance, 17 manifesting women have been reported in the literature^11,20,34–37^. Affected women usually manifest later in life than male patients and often present with a more parkinsonian phenotype. However, hyperkinetic movement disorders such as dystonia, chorea, and tremor have also been described in female XDP patients^11^. The genetic constellation reported in women manifesting XDP due to the SVA retrotransposon insertion includes homozygosity for the mutation in two cases^20,34^ or a typical or atypical Turner syndrome with X-chromosome monosomy in another reported patient^36^. In the present study, we included only women who did not report any symptoms and were therefore considered asymptomatic. Compared to HC, fMC had higher MDS-UPDRS III scores; however, the difference was not significant when corrected for higher AAE in the fMC group. Scores capturing dystonia (BFMDRS) or combined dystonia and parkinsonism (XDP-MDSP-RS) did not differ between fMC and HC.

In our previous TCS study of male patients with XDP, non-manifesting carriers, and HC, LN+ was more frequent in patients and was detectable in a subset of non-manifesting carriers several years before symptom onset^6^. In line with these findings, we also observed a higher proportion of LN+ in fMC compared with HC. Since clinical scores did not differ between women with and without LN+, LN+ itself does not appear to directly influence the clinical phenotype, although overall scores were low and did not indicate manifest disease. More sensitive approaches, such as gait and balance analysis using wearable sensors, may be better suited than conventional rating scales to detect subtle motor differences between groups^38^. However, both cDD and AAE were higher in fMC with LN+ than in those without LN+, whereas no such age effect was seen in HC, supporting the view that LN+ reflects underlying neurodegeneration. Taken together, LN+ can be regarded as an imaging phenotype of XDP in women. MRI-based quantification of LN properties may further refine this marker and facilitate its use as an imaging readout.

While LN+ is a characteristic TCS feature in atypical parkinsonian syndromes, increased substantia nigra echogenicity (SN+) is typically seen in idiopathic Parkinson’s disease (iPD). The finding that SN+ size was normal in fMC and did not differ from HC supports the notion that the parkinsonian signs in fMC are most likely driven by the XDP haplotype and striatal pathology rather than by coincident (prodromal) idiopathic PD.

Several mechanisms have been proposed to modulate the penetrance of XDP in women. Both homozygosity and X-chromosome monosomy were excluded in all participants in this study. Another mechanism that may contribute to phenotypic expression in heterozygous women is skewed XCI^11^. We did not find a significant influence of XCI on clinical scores or frequency of LN+. Notably, fMC with screwed XCI in favor of the wildtype allele, resulting in reduced expression of the mutated allele, showed no parkinsonian signs, and only one out of eight had LN+. Due to the small sample size and the possibility that XCI patterns may differ substantially between tissues such as blood and brain^39^, only cautious and limited conclusions can be drawn from these results. However, the question of whether screwed XCI might exert a protective effect warrants further investigation, ideally in post-mortem studies.

Estimates of PD prevalence in the Philippines vary. A nationwide questionnaire-based survey reported a prevalence of approximately 0.95% (950 per 100,000^7^, whereas a smaller community-based study combining questionnaires with case ascertainment reported a prevalence of roughly 0.55% (550 per 100,000)^40^. Notably, in contrast to most populations where PD is more common in men^41^, PD appears to be more prevalent among women in the Philippines^7^. It has been proposed that Filipino women with parkinsonism may be underdiagnosed with regard to XDP carrier status, which could contribute to this sex difference^11^. However, the prevalence of XDP in the general Philippine population is estimated at 0.34 per 100,000, with higher rates on Panay Island (5.25 per 100,000) and particularly in Capiz province (18.9 per 100,000)^42^. Given these prevalence figures and the subtle clinical effects observed in our cohort, it seems unlikely that parkinsonian fMC accounts for a substantial portion of the reported sex difference in PD prevalence. Nevertheless, genetic testing for XDP in women of Filipino descent presenting with parkinsonism could help clarify this issue.

Future studies are needed to elucidate the mechanisms underlying LN+, including post-mortem work and multimodal MRI studies linking structural imaging to ultrasound findings. Longitudinal investigations will be particularly important to define the temporal relationship between the emergence of LN+ and the onset of parkinsonian signs. Moreover, assessing additional phenotypic domains, such as cognitive function, will contribute to a more comprehensive understanding of XDP expression in women.

In conclusion, our findings indicate that women carrying the XDP haplotype frequently display LN+, highlighting LN+ as a relevant imaging marker of early neurodegenerative change in this population, especially in those with higher genetic risk.

## Acknowledgments

We thank all participants and the caregivers of patients with XDP for their participation and the administration of the Makati Medical Center for their valuable support. We thank Madita Grümmer for her work in study coordination and Pauline Tiegs for the support in data curation. Perplexity Pro (Perplexity AI) was used as a language and style assistant. The study was supported by intramural funding from the University of Lübeck to M.B. and H.H., funding from the German Research Foundation to C.K. and N.B. (FOR2488) and the Collaborative Center for X-Linked Dystonia-Parkinsonism (CCXDP) to C.C.D. and N.B. The authors MGP and MBo gratefully acknowledges support from the Detlef Zillikens Clinician Scientist Academy of Precision Health in Schleswig-Holstein (PHSH) and the Clinician Scientist Academy [Lübeck] The author T.K would like to acknowledge support from the Global Parkinson’s Genetics Program (GP2) funded by the Aligning Science Across Parkinson’s initiative and implemented by The Michael J. Fox Foundation.

## Authors Contribution

MGP and NB contributed to the conception and design of the manuscript; MGP, CCD, PC, MBo, JO, TK, JU, JQL, MBr, SMA, CK, AW and NB contributed to the acquisition and analysis of data; MGP, MBr, AW and NB contributed to drafting the text; MGP and NB contributed to preparing the figures.

## Data availability

The datasets generated during the current study are not publicly available due to the presence of individual genetic and clinical information that could compromise participant privacy, but are available from the corresponding author on reasonable request.

